# Histological examination of choroid plexus epithelia changes in schizophrenia

**DOI:** 10.1101/2023.03.10.23286921

**Authors:** MR Williams, CM Macdonald, F Turkheimer

## Abstract

**Background:** The choroid plexus (CP) is suggested to be involved in neuroimmune regulation avia the interaction between central and peripheral inflammation. Quantitative imaging has demonstrated volumetric CP change in psychosis, schizophrenia and depression. This study histologically examines CP epithelial cell morphology in these illnesses to identify the biological source of such volumetric changes.

**Methods:** Formalin-fixed paraffin-embedded FFPE blocks were obtained bilaterally from the lateral ventricles of 13 cases of sex- and age-matched brains from each of schizophrenia (SZ) with psychosis, major depressive disorder (MDD) and matched controls. FFPE blocks were sectioned at 7μm and routinely stained for H&E. Morphological analysis of 180 CP epithelia/case was conducted blindly on digital images collected x600 magnification. Calcification was assessed in all CP regions manually.

**Results:** Linear General Model analysis shows a significant effect of diagnosis on somal width (p=0.006, r^2^=0.33, adj=0.25), with a significant difference between SZ non-medicated v non-medicated NPD controls (p=0.032), demonstrating increased somal width in SZ with psychotic medication but not in unmedicated SZ cases. No effects were observed in calcification.

**Discussion:** Epithelial cells examined were adhered to CP fibrous surface so width expansion describes the primary methods for these cells to expand with adherence to this surface. The interaction of antipsychotic medication and diagnosis demonstrates that this is an illness-specific change and mediated through the DA-system, although the mechanism is unclear at present.

## Introduction

The choroid plexus (CP) has been identified as a distinct structure for many years and investigated for its possible role in the pathophysiology of schizophrenia (SZ) for over a century^1-3^. The CP is important for the secretion of cerebrospinal fluid (CSF). It is formed of a vascularized stroma with loose connective tissue with a single outer layer of epithelia continuous with the ependyma lining the brain of lateral, 3rd and 4th ventricles, as well as often found on the outer surface of the brain^4-6^. The CP is suggested to be regulated by descending neurons and by circulating factors. High receptor concentrations for circulating hormones and serotonin, atrial natriuretic peptide, vasopressin and the insulin-like growth factors have been reported in the CP and shown to affect its function^7^.

The CNS is protected against substances from the blood by the blood–brain barrier (BBB) and the blood–cerebrospinal barrier^8-11^. CP epithelial cells form the blood–cerebrospinal fluid barrier, controlling CSF secretion into the ventricular system^12^. The production of CSF from blood plasma in the CP is performed by a filtration process to remove unneeded plasma proteins, controlling CSF protein content^8,13^. The blood–cerebrospinal barrier has a key role in the spreading of inflammatory reactions from the peripheral regions to the CNS and has been suggested to contribute to the pathogenesis and progression of various neurological disorders^14-16^, demonstrated by the report that up to 40% of patients with inflammatory bowel disease present with psychosocial disturbances^17^.

Recently, CP imaging abnormalities have been reported in psychiatric cohorts. In patients with SZ, CP volume has been found consistently larger compared to controls^18-20^, whereas the enlargement has demonstrated a genetic component as it was intermediate in first-degree relatives^18^. This finding has been repeated trans-diagnostically as greater CP volume has also been demonstrated in patients with Major Depression Disorder (MDD)^20^. There is evidence of CP enlargement with measures of inflammation, either peripheral or central, and stress. In patients with SZ - a positive association has been demonstrated with peripheral plasma IL-6 levels and allostatic load^18,19^. In MDD, CP volume has been associated with neuroinflammation measured with positron emission tomography (PET) in the anterior cingulate cortex, prefrontal cortex and insular cortex, but not with peripheral inflammatory markers^20^. Notably CP enlargement has been recently proposed as a transdiagnostic marker of brain inflammation in neurodegenerative conditions^21^.

Imaging studies in-vivo are limited though as they may not generally indicated the cellular bases of CP volumetric changes. A small CT-MRI study of 12 SZ patients under 45y has implicated calcification in the CP volumetric changes following neuroleptic treatment^22,23^. Calcification of the CP as measured by quantitative imaging has been implicated in serotonin function in SZ and as a marker of depression^24,25^.

The aim of this neuropathological study was to verify the cellular abnormalities associated with CP volumetric changes in SZ and MDD cohorts. Specifically, we apply digital histopathological methods to examine the morphology of epithelial cells in CP from the lateral ventricles, and manual examination of CP calcification from multiple brain areas from human neuropathological samples from control, SZ with psychosis and MDD cases.

## Methods

### Cases

Cases were sourced from the Corsellis brain collection, selected for age of death between 45-60 and re-reviewed for inclusion by modern criteria as defined by ICD-10 by a senior consultant psychiatrist. SZ cases all contained first-rank symptoms with onset below 30 years, with positive symptomatology including psychosis with auditory hallucinations. MDD cases were included with repeated cycles of treatment-resistant depression with onset under 25 years. Cases were included only if they had substantial tissue available for use with full clinical and psychiatric files with drug histories and passed neuropathological review to exclude confounding pathology. A breakdown of cases included in this study is shown in Table 1.

**Table 1.** NPD- no psychiatric disorder, SZ- schizophrenia, MDD- major depressive disorder, NK- not known.

| Case | Age/y | Sex | Diagnosis | PM/h | Fix/y | Suicide | Cause of death |
| --- | --- | --- | --- | --- | --- | --- | --- |
| 1 | 51-55 | M | NPD | 82 | 38 | N | Heart Failure from COPD |
| 2 | 45-50 | F | NPD | 23 | 30 | N | Non-follicular lymphoma |
| 3 | 45-50 | M | NPD | 61 | 39 | N | Lymphoid leukaemia |
| 4 | 56-60 | F | NPD | 98 | 26 | N | Chronic ischaemic heart disease |
| 5 | 51-55 | M | NPD | 24 | 29 | N | Congestive heart failure |
| 6 | 51-55 | F | NPD | 36 | 35 | N | Malignant neoplasm of stomach |
| 7 | 56-60 | F | NPD | NK | 40 | N | Pneumonia |
| 8 | 51-55 | F | NPD | 54 | 45 | N | Acute myocardial infarction |
| 9 | 56-60 | F | NPD | 60 | 40 | N | Pneumonia |
| 10 | 51-55 | M | NPD | 56 | 39 | N | Acute myocardial infarction |
| 11 | 45-50 | M | NPD | 32 | 37 | N | Acute myocardial infarction |
| 12 | 56-60 | M | NPD | 55 | 40 | N | Atherosclerosis |
| 13 | 51-55 | F | NPD | 24 | 41 | N | Gastrointestinal haemorrhage |
| 14 | 56-60 | F | SZ | NK | 51 | N | Pneumonia |
| 15 | 45-50 | M | SZ | 46 | 26 | Y | Suicide |
| 16 | 56-60 | F | SZ | 22 | 39 | N | Pulmonary embolism |
| 17 | 56-60 | M | SZ | 28 | 38 | N | Congestive heart failure |
| 18 | 51-55 | F | SZ | 28 | 46 | N | Pulmonary embolism |
| 19 | 56-60 | M | SZ | 58 | 47 | N | Post-operative complications |
| 20 | 56-60 | M | SZ | 24 | 41 | N | Acute myocardial infarction |
| 21 | 56-60 | F | SZ | 26 | 38 | N | Congestive heart failure |
| 22 | 45-50 | M | SZ | 91 | 47 | N | Congestive heart failure |
| 23 | 45-50 | M | SZ | 29 | 47 | N | Acute myocardial infarction |
| 24 | 56-60 | F | SZ | 19 | 40 | Y | Intentional self-harm by hanging |
| 25 | 56-60 | F | SZ | 26 | 42 | N | Acute myocardial infarction |
| 26 | 56-60 | F | SZ | 41 | 48 | N | Kidney Failure |
| 27 | 45-50 | F | MDD | 66 | 30 | Y | Intentional self-poisoning |
| 28 | 51-55 | F | MDD | 63 | 27 | Y | Intentional self-poisoning |
| 29 | 45-50 | M | MDD | 26 | 24 | N | Asphyxiation |
| 30 | 51-55 | F | MDD | 22 | 28 | Y | Suicide |
| 31 | 56-60 | M | MDD | NK | 31 | N | Pulmonary embolism |
| 32 | 45-50 | M | MDD | 17 | 30 | Y | Intentional CO poisoning |
| 33 | 45-50 | F | MDD | 28 | 31 | Y | Intentional self-poisoning |
| 34 | 45-50 | M | MDD | 16 | 25 | Y | Intentional CO poisoning |
| 35 | 56-60 | M | MDD | NK | 33 | Y | Intentional self-harm by jumping |
| 36 | 51-55 | M | MDD | 73 | 35 | Y | Intentional self-harm by jumping |
| 37 | 56-60 | F | MDD | NK | 33 | N | Pneumonia |
| 38 | 56-60 | F | MDD | 52 | 35 | N | Acute myocardial infarction |
| 39 | 56-60 | M | MDD | 21 | 38 | N | Pneumonia |

### Tissue source and processing

The tissue was originally from the Corsellis brain collection, formerly the Runwell collection^26^, currently held at the Gordon Pathology Museum (Guys Campus, KCL, London, UK). Dissection was conducted in the museum processing rooms consistent with previous neuropathology projects^27-32^. CP tissue was dissected bilaterally from the lateral ventricles of cases described in Table 1. Right and left lateral ventricles were found within coronal sections and removed separately for each ventricle, stored in a single pathology cassette for each case, with dissection markers to distinguish laterality. Bilateral CP samples were obtained from all cases. Dissected tissue was contained in standard pathology cassettes and transported in 10% formalin (4% v/v).

Tissue processing, sectioning and staining was conducted by Histologix Ltd (Nottingham, UK) using routine methods and embedded into paraffin blocks. Formlin-Fixed Paraffin-Embedded (FFPE) blocks were sectioned by microtome at 7μm and mounted on electrostatic 75×25mm slides. Routine H&E staining was performed on all sections with Harris Hematoxylin and 50/50 aqueous Eosin using a Leica ST4040 linear slide stainer (Leica, Germany).

### Image collection

Images were collected on a single Olympus Microtec microscope (Tokyo, Japan) at x600 total magnification using a 10MP USB 2.0 Colour CMOS C-Mount Microscope Camera (Amscope, USA) at 3584×2748 resolution.

Initially CP tissue was identified at x150 magnification. For each image a grid mask was imposed and a random number generator (www.calculator.net) used to select sampling regions to remove selection bias.

Regions containing >30 measurable epithelia, as defined by inclusion criteria, were taken from each hemisphere per slide to give 2 regions sampled per slide. This was conducted on 3 slides per case to give a total of 6 regions containing 180 measured cells per case.

Experimental measures were made using a rubber stylus and touchscreen interface on an IPad Air2 device using the Segmentum Imaging v1.1 App (Segmentum Analysis Ltd, UK). This was done through serial measurement of graticule-calibrated whole image analysis using ruler and region functions for measurement of distances and circumferences/areas respectively. Data was collected blind to case identification.

### Assays and measurements

Epithelial cells for measurement were identified from images captured as described. CP cells for measures were defined as “surface-based epithelial cells attached to the basal substrate with clear membranes and visible nucleus”, shown in Fig 2.

**Fig. 1.**
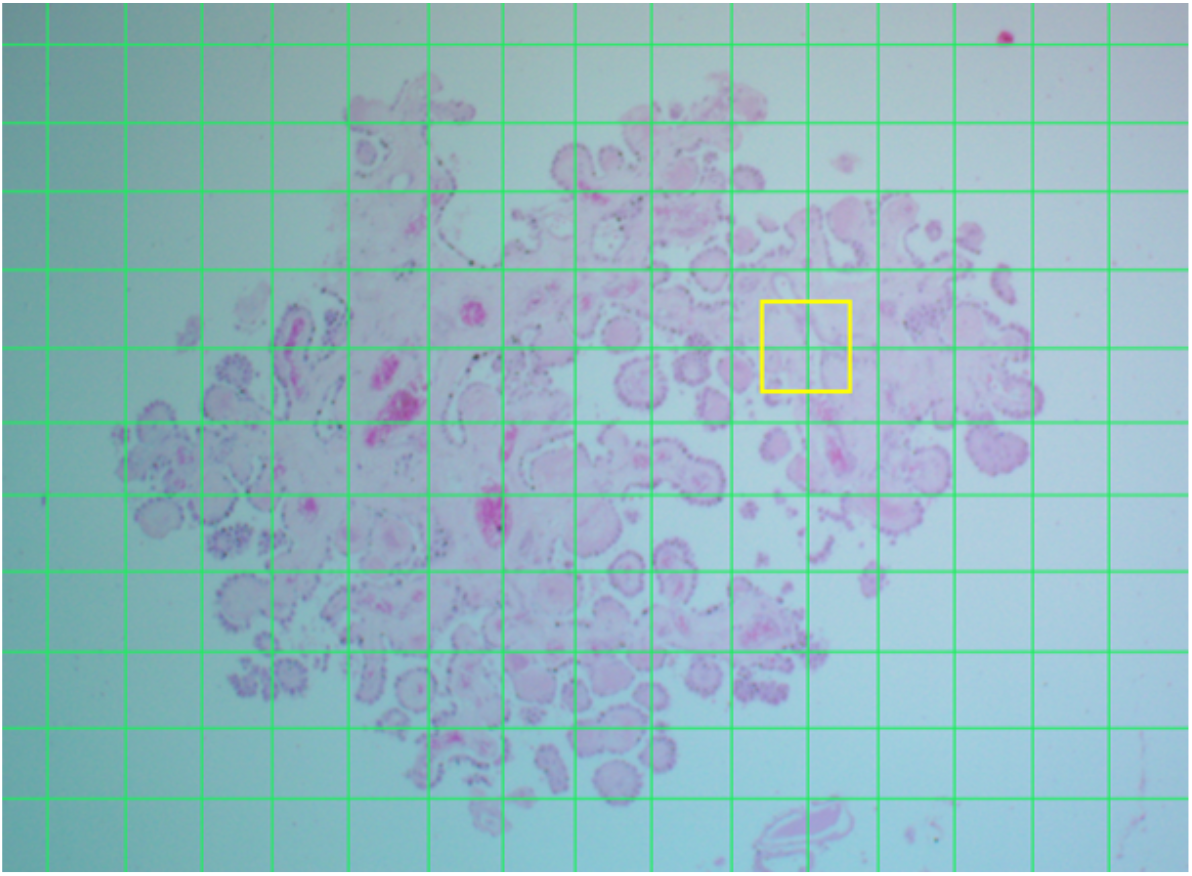
H&E-stained CP at x150 magnification with grid mask (Green). Yellow square indicates region taken at x600 magnification for measurement.

**Fig. 2.**
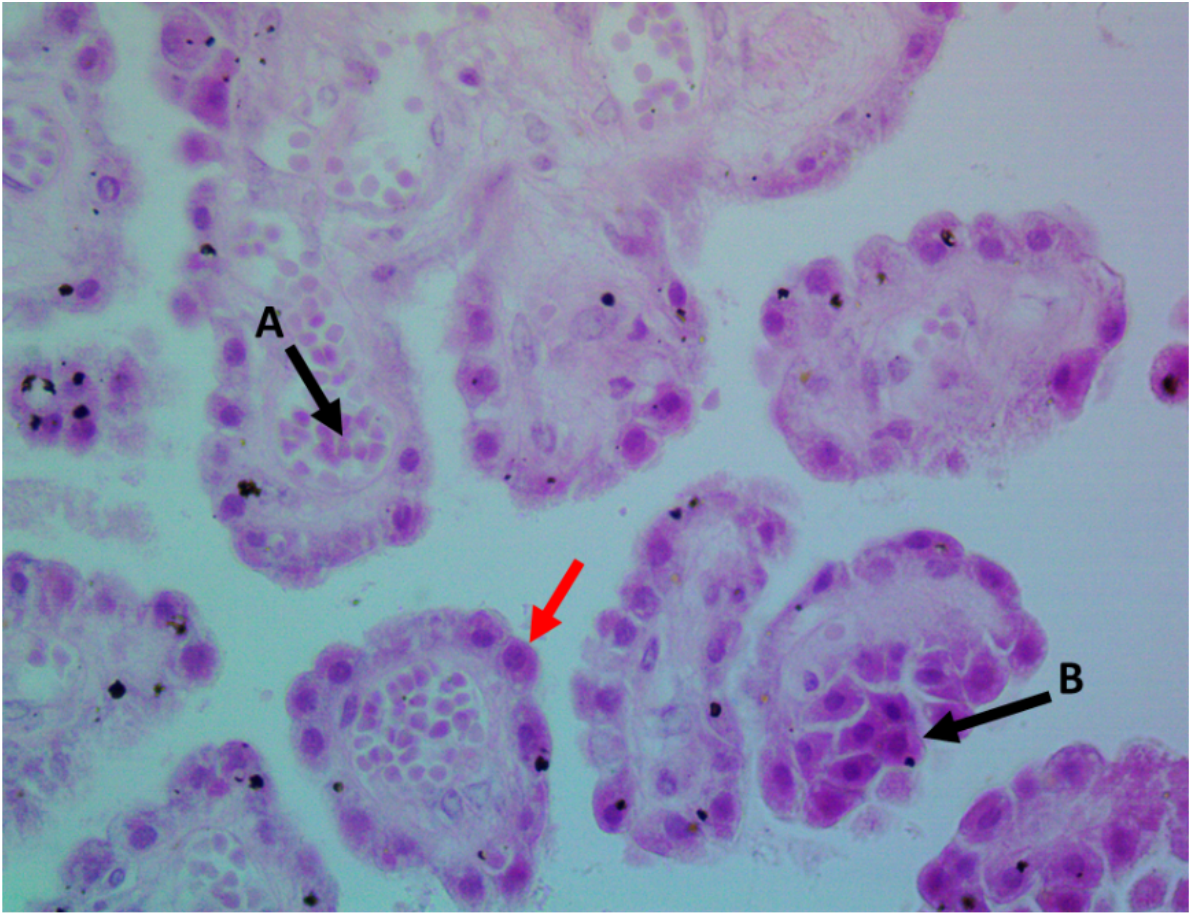
H&E-stained CP at x600 magnification. Red arrow indicates epithelial cell for measurement. A – erythrocytes in blood vessel, B – epithelial cell not included for measurement due to not being attached to the basal substrate.

Repeated measures assays were conducted on 67 CP epithelia encompassing two randomly selected cases to determine statistically significant *n* size. Measures tested were:

1. Cell length - defined as the longest distance between the basal surface and the opposing cell membrane.
2. Cell width - defined as the longest distance between cell membrane surfaces not connected to the basal surface.
3. Cell circumference - defined as the length of the continuous outer membrane.
4. Cell cross sectional area - defined as the measured area of the cell bound by its outer membrane.
5. Nucleus length - defined as the distance of the cell nucleus at its longest axis.
6. Nucleus width - defined as the widest point of the nucleus perpendicular to the nucleus length.
7. Nucleus circumference - defined as the length of the outer surface of the nucleus.
8. Nuclear cross-sectional area - defined as the measured area of the nucleus bound by its outer membrane.

Somal measures on an example epithelial cell are described in Fig. 3. Nuclear measures were conducted in a similar fashion.

**Fig. 3.**
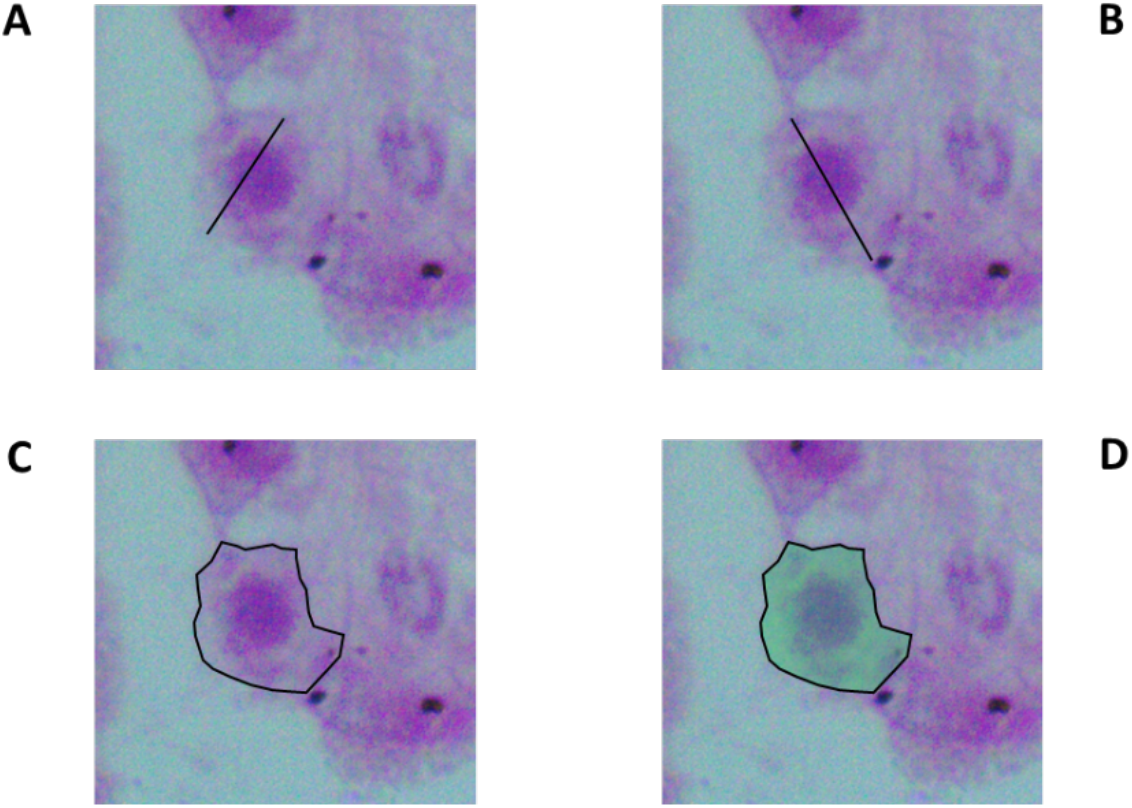
H&E-stained CP at x600 magnification. Red arrow indicates epithelial cell for measurement. A – Somal length, B – Somal width, C - Somal circumference, D – Somal cross sectional area.

Repeated measures demonstrated that the appropriate minimum sample size was 30 cells measured per image to meet the required *n* size for all scalar measures (Table 2).

**Table 2.** Ideal n sizes for each of the measures. NA – Not Applicable.

| Measure | Ideal $n$ size |
| --- | --- |
| Cell length | 30 |
| Cell width | 25 |
| Cell circumference | 25 |
| Cell CSA | NA |
| Nucleus length | 20 |
| Nucleus width | 20 |
| Nucleus circumference | 30 |
| Nucleus CSA | NA |

Experimentally each case involved measurement of 30 cells per image, 3 images per hemisphere, bilateral measures giving 6 images per slide. This resulted in the measurement of 180 cells per case by methods consistent with previous pathology studies^28,29,32^.

Intra-rator errors were <5% for all measures.

### Calcification

Calcification of CP in various ventricular regions was assessed semi-quantitatively. The areas of CP analysed were right and left lateral ventricles, 3^rd^ ventricle, cerebral aqueduct, surrounding surfaces of the brainstem and pons and the outer surface of the frontal lobe.

CP was not found in one region of each of 4 cases (11/91, 107/86, 48/88, 31/90). These were not the same regions absent in any case. Tissue was manually assessed whilst in formalin, with CP in each area manually rated as having no calcification, a small amount of calcification, a significant amount of calcification or was completely calcified.

### Statistical analysis

Measures were first tested using a general linear model (GLM) where independent factors were diagnostic group, age and antipsychotic use. Given the number (eight) of different measures used multiple testing correction was considered and performed via the Bonferroni correction. Only for those measures where the GLM was significant (p<0.05 corrected), post-hoc F tests were then performed based on linearly independent pairwise comparisons among the estimated marginal means.

Statistical analysis was run using SPSS v28.0.0.1 (IBM Corp).

### Ethics

This project was conducted under ethical permission from Brain UK review number 003/12.

## Results

Linear General Model analysis showed a significant effect of diagnosis on somal width (*p*=0.006, r^2^=0.33, adj=0.25) that survived Bonferroni correction. There was a significant difference between SZ non-medicated v non-medicated NPD controls (*p*=0.032m Fig. 4), demonstrating increased somal width in SZ with psychotic medication but not in unmedicated SZ cases. Age was also positively associated with somal width (*p*=0.048).

**Fig. 4.**
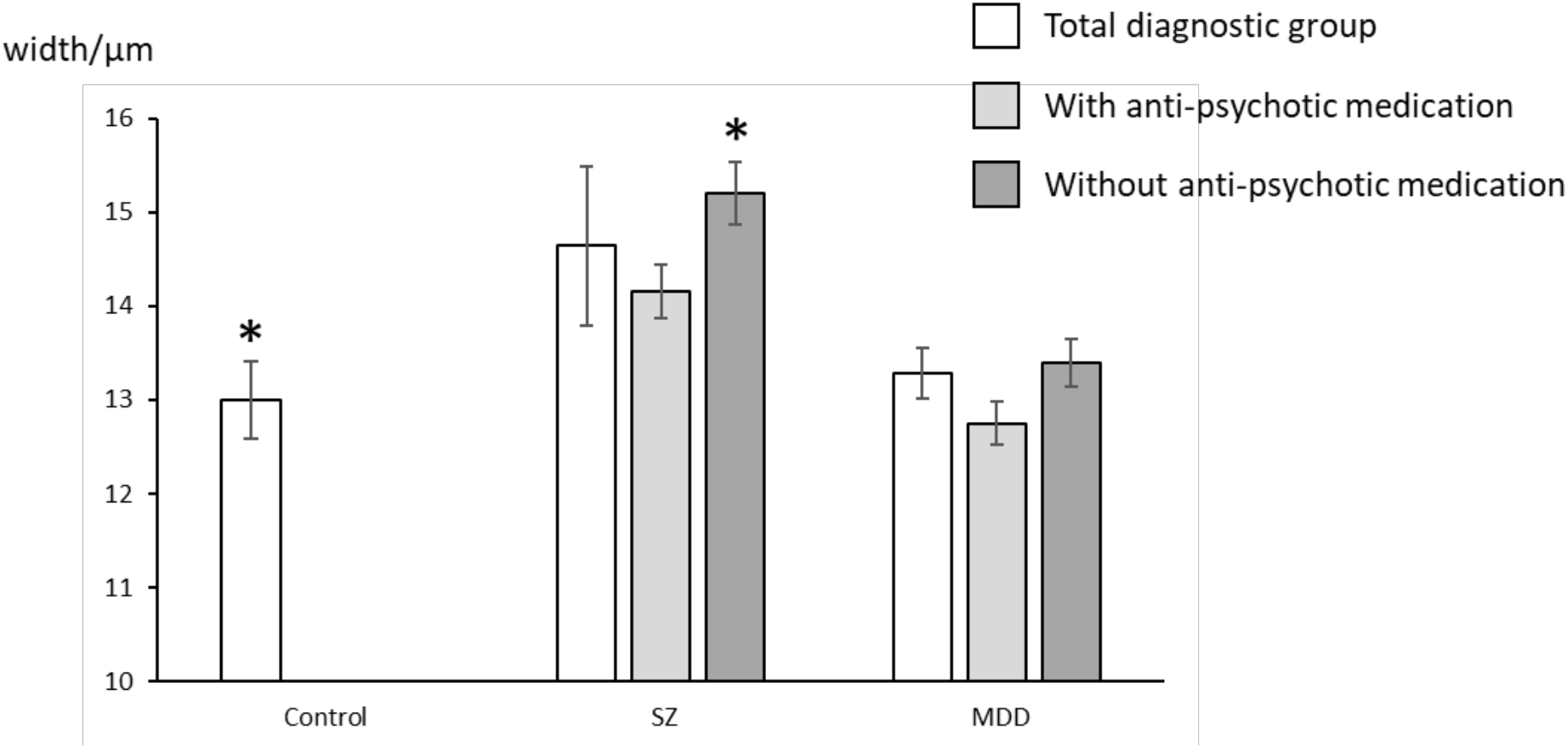
Somal width by diagnosis and anti-psychotic treatment. *represents significant difference between controls and SZ without medication (LGM, *p*=0.032). Data shown as mean ± SEM.

No significant associations with independent factors were observed in somal length, circumference or cross-sectional area, and no changes were observed in nuclear morphology, and no effects were observed in calcification (data shown in Tables 3 & 4).

**Table 3.**
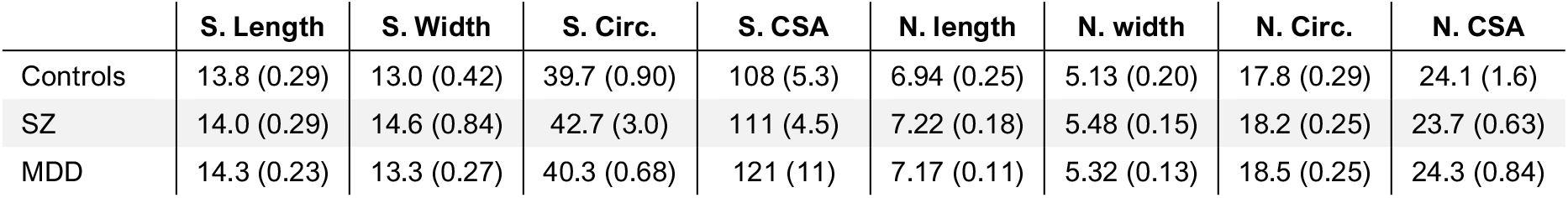
Means of measures for diagnostic groups. S – Somal, N – Nuclear, CSA - Cross Sectional Area. Data shown as μm, except for CSA results shown as μm^2^. Data shown as mean (3sf) ± SEM to (2sf).

**Table 4.** Calcification results by diagnostic group. RLV – Right Lateral Ventricles, LLV – Left Lateral Ventricle, 3^rd^ V – Third Ventricle, 4^th^ V – Fourth Ventricle, Cer Out – Outer surface around cerebellum, Ctx Out – Outer surface of frontal lobe cortex. NB: 4^th^ ventricle describes the area around the upper part of the brainstem distinct from the cerebellum also. Data shown as mean ± SEM to 2sf.

|  | RLV | LLV | 3<sup>rd</sup> V. | 4<sup>th</sup> V. | Cer. Out. | Ctx. Out. |
| --- | --- | --- | --- | --- | --- | --- |
| Controls | 0.31 (0.13) | 0.45 (0.18) | 0.69 (0.26) | 0.38 (0.18) | 0.15 (0.10) | 0.53 (0.22) |
| SZ | 0.31 (0.17) | 0.38 (0.18) | 0.77 (0.32) | 0.38 (0.21) | 0.15 (0.10) | 0.53 (0.22) |
| MDD | 0.31 (0.13) | 0.54 (0.24) | 0.70 (0.29) | 0.50 (0.19) | 0.30 (0.17) | 0.30 (0.21) |

## Discussion

The main finding of this work is a significant increase in the somal width of epithelial cells in the CP of SZ cases. The epithelial cells examined were adhered to the CP fibrous surface, therefore width expansion is the primary method for expansion a cell to expands and has multiple possible causes. It could originate from changes between the cells and the blood capillaries, changes in the gap and tight junctions between the cells themselves, from the altering of their structural organisation, or from within the cells themselves. The cause of such changes could be from descending neural projections, endocrine routes or perhaps from other peripheral sources.

The interaction of antipsychotic medication and diagnosis demonstrates that this is an illness-specific change and mediated through the DA-system, although the mechanism is unclear at present. Rat CP contains DA receptor proteins, with D1 receptors predominantly expressed in the epithelium and the arterial smooth muscles and D2 receptors expressed in sympathetic fibres innerving CP epithelium and vasculature^33^. DA infusion has been demonstrated to increase blood flow through the CP in the brain of the sheep^34^, and in mice DA genetic-hypofunction reduces CP permeability^35^. This finding adds to previous evidence in pre-clinical models on dopaminergic modulation of CP functionality as a promising therapeutic strategy for neurodevelopmental and psychiatric disorders.

Although we were able to demonstrate CP volumetric alterations in the brains of subjects with SZ, we did not find such alterations in MDD patients, although increased CP volume has been demonstrated by imaging in an MDD cohort that largely presented with heightened peripheral immunity and such state may not be represented in the cohort used here for which allostatic load was not available^22^.

Methodologically, measurement of epithelial CP has several difficulties to overcome due to the nature of the tissue. The cell organisation is likely the reason behind the very high number of cells that had to be measured by case to get useful results. Nucleoli measures could not be conducted due to the rare visibility of nucleoli in the epithelia present, with it being unusual to even see one clear nucleolus in 30 nuclear assessments in an image.

If further assays could be developed it may be possible to quantitatively assess the calcification in these samples microscopically, although given the null macroscopic result presented and the rare occurrence on the slides this would be of questionable value.

Further pathological examination of CP tissue for GFAP or S100β as astrocyte markers may yield insights in to CP disruption, as astrocytes are known to be affected across wide regions of the brain in SZ^28,30,36-41^, and have a particular role in inflammation^42,43^, having a role in CP structure and function as in the CNS^44,45^.

Evans blue stain has been successfully used to examine BBB integrity in live rodents^46,47^, and has been successfully used to identify regions of BBB damage by injection of an Evans Blue-Hoechst cocktail for pathological analysis^48^. It is still used *in vivo* for medical diagnostics^49^. Evans Blue has also been demonstrated to be a marker of cell death or damage in plants^50-52^. Although plant cells are notably different from animal cells, the Evans Blue stain identifies such damage by membrane permeability, which may have value in translation to animal pathology.

## Data Availability

All data produced in the present study are available upon reasonable request to the authors

## Acknowledgements

This project was funded by the funded by the NIHR Maudsley Biomedical Research Centre at South London and Maudsley NHS Foundation Trust and King’s College London.

Tissue samples were obtained from Guy’s and St Thomas’ NHS Foundation Trust as part of BRAIN UK, supported by Brain Tumour Research, the British Neuropathological Society and the Medical Research Council, Ethics ref: Brain UK 12/003 - Neuropathological examination of neurons, glial cells, axons and molecular factors in mood and affective disorders^53^.

The authors would like to acknowledge the support of the Mr. William Edwards Gordon Pathology Museum at Kings College London Bridge campus for his care of the tissue and support of dissection, storage and transport.

Dr. Williams is inventor of Segmentum Imaging, the pathology software used in this study, and director of the Segmentum Analysis. This software was provided free for this study.

